# Comparison of DXA and HR-pQCT Measures Among Female Runners at Risk for Relative Energy Deficiency in Sport (REDs): A Pilot Study

**DOI:** 10.1101/2025.09.08.25335357

**Authors:** Morgan Smith, Julia Flora, Kyla Kent, Jin Long, Yutong Zhong, Aubrey Roberts, Michael P. Snyder, Emily Kraus

## Abstract

**Objectives:** Relative Energy Deficiency in Sport (REDs) is a syndrome driven by problematic low energy availability, impairing physiological and/or psychological function. While REDs assessment typically utilizes dual energy X-ray absorptiometry (DXA) of areal bone mineral density (BMD), high-resolution peripheral quantitative computed tomography (HR-pQCT) offers additional insight into bone microarchitecture, geometry, and volumetric BMD. This study aimed to evaluate bone health among female runners at risk for REDs using DXA and HR-pQCT measures.

**Methods:** Female runners aged 18-30 years, training at least 5 hours/week, were recruited and underwent anthropometric measurements, VO_2_ max testing, clinical laboratories, DXA scans, and HR-pQCT imaging of the tibia. REDs risk was assessed using validated questionnaires, clinical laboratories, and physician interviews. Participants were categorized as no-risk (green) or at-risk (yellow/orange/red) for REDs.

**Results:** Twenty-one participants (age 26 ± 3 years) completed the study. Six were classified as no-risk and fifteen as at-risk for REDs. Sub-clinically low BMD (z-score ≤-1) was the most prevalent indicator in at-risk participants. The at-risk group reported significantly higher weekly mileage (>40 miles/week: 66.7% vs 33.3%, p=0.043) and lower maximum extensor strength on muscular endurance testing (p=0.015). While no other between-group differences reached statistical significance, 12 of 14 HR-pQCT values showed poorer outcomes in the at-risk group.

**Conclusions:** Despite the small sample size, this pilot revealed consistent HR-pQCT trends suggesting potential links between REDs risk and compromised bone geometry, microarchitecture, and volumetric BMD. Integration of HR-pQCT with REDs screening may provide a more comprehensive characterization of bone health compared to DXA alone.

**SUMMARY BOX:** *What is already known on this topic:* Relative Energy Deficiency in Sport (REDs) is a clinical syndrome driven by problematic low energy availability that negatively impacts bone density and increases risk for bone stress injury (BSI).

*What this study adds:* Consistent trends in HR-pQCT outcomes demonstrate possible patterns of compromised bone geometry, microarchitecture, and volumetric bone mineral density in athletes with elevated REDs risk.

*How this study might affect research, practice or policy:* Our findings suggest that integrating HR-pQCT imaging with REDs screening may provide a more comprehensive characterization of REDs-related bone health than DXA alone. Further research examining the relationship between REDs risk and detailed bone health measures, including geometry and microarchitecture, is warranted to better inform clinical practice and screening tools.

## BACKGROUND

Problematic low energy availability (LEA), defined as a state where energy expenditure exceeds energy intake over a prolonged period, has been identified as a contributor to compromised bone health. LEA and its associated symptoms, including secondary amenorrhea (absence of period for three or more months in a row) and hormonal dysfunction, can contribute to an increased risk of low bone mineral density (BMD) [1,2]. Compromised bone health characterized by low BMD, altered geometry, and deficits in microarchitecture may serve as important predictors of fracture risk [3–9].

LEA is highly prevalent in female athletes and is the underlying mechanism of Relative Energy Deficiency in Sport (REDs) [10,11]. REDs was defined by the International Olympic Committee (IOC) in 2014 as a clinical syndrome caused by insufficient energy intake that results in impaired physiological and/or psychological function [12]. REDs-related LEA has been shown to negatively impact bone density and remodelling, and increase fracture risk [13]. In 2023, the IOC refined the diagnostic process by introducing the REDs Clinical Assessment Tool 2 (CAT2) [14], an evidence-based tool for evaluating athletes’ risk for REDs. The REDs CAT2 assesses severe primary, primary, and secondary indicators to classify athletes into a green, yellow, orange, or red risk category. This classification is based on the quantity and severity of indicators present; the more indicators present, the higher the risk level.

One of the main quantitative variables included in the REDs CAT2 is areal BMD z-score assessed by dual-energy X-ray absorptiometry (DXA). DXA’s accuracy is impacted by body size and composition, and it does not measure true volumetric bone mineral density or distinguish between trabecular and cortical compartments. High-resolution peripheral quantitative computed tomography (HR-pQCT) provides true volumetric bone mineral density (vBMD) for total, trabecular, and cortical bone. It also captures bone geometry, such as cortical area and thickness, and microarchitecture, including trabecular spacing, tissue mineral density, and cortical porosity. These parameters have been shown to be important predictors of fracture risk and overall bone strength, particularly in the tibia [8,9,15]. Additionally, DXA does not assess bone geometry or microarchitecture [16–18], providing stronger support for including the HR-pQCT measurement.

Few studies have evaluated HR-pQCT measures within individuals at risk of REDs [19], and a comparison of both DXA and HR-pQCT within a cohort of at-risk female runners has been lacking. Several studies have more broadly examined the relationship between LEA or amenorrhea and bone microarchitecture, reporting that athletes with either condition tend to have poorer bone outcomes [20]. The primary aim of this study was to evaluate bone health in female runners at varying risk levels for REDs, using DXA at the lumbar spine, proximal femur, and whole body to assess areal bone density, and HR-pQCT at the tibia to assess metaphyseal and diaphyseal bone geometry, microarchitecture, and volumetric density. We hypothesised that athletes at risk for REDs would demonstrate lower bone mineral density across both modalities, poorer microarchitecture, and altered tibial geometry compared with those not at risk.

## METHODS

### Ethical Approval, Participant Selection, and Screening

We identified and recruited 26 females from the San Francisco Bay Area, California, through flyers, local running groups, social media posts, orthopaedic sports medicine clinics, and snowball recruitment through referral. The following inclusion criteria were used: i) female, ii) 18-30 years of age, iii) run training at least 5 hours per week, iv) actively competing in either track, cross country, triathlon, or running as the majority of the training for their sport, and v) have zero to all of the following criteria: at least one bone stress injury in the past two years, eating disorder diagnosis or treatment within the past two years, abnormal periods, and a subjective decrease in performance in the last three months. Abnormal periods were defined as the absence of three or more periods in the last 12 months. Criteria (v) was used to screen potential level of risk for REDs to monitor recruitment of risk levels and attempt to recruit a balanced sample. The following exclusion criteria were used: i) pregnancy or lactation in the past 12 months, ii) planning to become pregnant in the next 4 months, and iii) hormonal contraceptive use. This pilot study was conducted with approval from the Institutional Review Board of Stanford University (IRB 69308), the Student Athlete Research Oversight Committee, and the Student Data Oversight Committee. Interested participants were screened for eligibility, consented, and oriented to the study in a 30-minute meeting, conducted either in person or via Zoom. All participants signed written informed consent prior to participating.

#### GENERAL STUDY DESIGN

Once consented, participants were provided with ovulation kits and instructions for menstrual cycle tracking, along with secure links to complete online questionnaires. Participants were then scheduled for two visits on separate days, in no particular order: i) main study visit that included all anthropometric, DXA, HR-pQCT, and muscle strength assessments by Biodex, and ii) a hospital visit for clinical laboratories and VO_2_ max testing. Ovulation testing, menstrual cycle tracking, and weekly surveys began with each participant’s first menstrual period following consent and continued throughout a three-month period. Study data were collected and managed using REDCap electronic data capture tools hosted at Stanford University [21,22].

#### SPECIFIC TESTING PROCEDURES

##### Ovulation Testing and Menstrual Cycle Tracking

Each participant was provided with 50 Easy@Home (by Premom) luteinizing hormone (LH) test strips. Eumenorrheic participants were instructed to begin testing their first morning urine sample starting on the seventh day after the onset of menstrual bleeding. They were asked to continue daily testing until they observed a positive test result, and to report via a weekly survey through REDCap [23]. Participants also tracked positive ovulation tests and menstrual bleeding days and symptoms using the Clue period tracking app by Biowink GmbH.

##### Anthropometric Measurements

Participants underwent anthropometric measurements at the Stanford Assessments of Muscle and Bone across the Ages (SAMBA) Lab during the main study visit. Pregnancy tests were administered prior to assessments and imaging per standard protocol. Height and sitting height were measured in millimetres (mm) to the nearest 1 mm with a wall-mounted stadiometer, and weight was measured in kilograms (kg) to the nearest 0.1 kg using a digital scale. Tibia length was measured in millimetres using a measuring tape. Body mass index (BMI) was calculated as weight in kilograms divided by the square of height in meters (kg/m^2^).

##### Muscle Strength

Isokinetic knee extensor and flexor strength and endurance of the dominant leg (participant selected) were evaluated using a Biodex System 4 Pro (Biodex Medical System Inc, Shirley, NY, USA) dynamometer. Participants were seated in the dynamometer chair in a standardized position of 85° hip flexion from the anatomical position and stabilized with a lap belt. The lever arm was aligned with the lateral epicondyle of the knee. The tested thigh was stabilized with a belt, and the ankle was attached to the device (slightly above the medial malleolus). Range of motion was defined for 85° (knee flexion 5° to 90°). Strength was assessed by a set of 5 maximal effort repetitions at 60 degrees/second. The best effort was reported. Endurance is assessed by a set of 30 repetitions at 300 degrees/second at the highest speed possible.

##### Dual-Energy X-ray Absorptiometry (DXA)

Measurements of areal bone mineral density were obtained for the lumbar spine, proximal femur (total hip and femoral neck), and whole body using the Horizon A DXA System (Hologic Inc., Bedford, MA, USA) with Apex software v5.5 and standard positioning techniques. Whole body DXA scans also provide measures of body composition, including total lean mass and fat mass, appendicular lean mass, and indices of lean and fat mass.

##### High-Resolution Peripheral Quantitative Computed Tomography (HR-pQCT)

Relative offset scans were performed using HR-pQCT (XtremeCT II, Scanco Medical, Bruttisellen, Switzerland) at the metaphyseal and diaphyseal tibia of the dominant leg, with a protocol of a centred stack at 7.3% and 30% respectively. Scans were repeated up to two times if excessive motion was detected. Trabecular BMD was assessed exclusively at the diaphyseal tibia, while vBMD was measured at both sites for total and cortical compartments. Bone geometry measures of total and cortical area, as well as microarchitecture assessments of cortical thickness, tissue mineral density, and cortical porosity were also performed at both sites. Additional microarchitectural measures, including bone volume fraction, trabecular number, trabecular thickness, and trabecular spacing, were evaluated at the metaphyseal site only.

##### REDs Risk Assessment and Caloric Intake

Participants completed a training and health history questionnaire and validated surveys, including the Low Energy Availability in Females Questionnaire (LEAF-Q), Eating Disorder Examination Questionnaire (EDE-Q), and Exercise Dependence Scale Questionnaire (EDS-Q). Fasted (for 8 hours) morning clinical laboratories (cholesterol panel and triiodothyronine (free T3)) from the hospital visit were used to inform risk level assessment. Participants also completed the Diet History Questionnaire III (DHQ-III) to determine daily energy intake. Following the completion of the questionnaires, physician-led interviews were conducted privately and virtually to clarify survey answers, confirm REDs risk level categorization, and screen for emerging indicators of REDs, including iron deficiency and urinary incontinence. All information provided via survey or interview was self-reported and was not validated with participants’ medical records. Risk levels for REDs (green, yellow, orange, or red, with the red risk level indicating highest risk) were determined using the criteria listed in Table 1 using the REDs-CAT2 [14]. Due to sample size limitations and previous literature [19,24], participants were grouped into no-risk (green) and at-risk (yellow, orange, or red) groups based on their REDs risk level.

**Table 1.** Distribution of REDs Risk Factors.

|  | All (n=21) | No-Risk (n=6) | At-Risk (n=15) |
| --- | --- | --- | --- |
| **Severe Primary Indicator** | **Number (%)** | **Number (%)** | **Number (%)** |
| Primary amenorrhea<sup>a</sup> | 3 (14.3%) | 0 (0.0%) | 3 (20.0%) |
| **Primary Indicators** | **Number (%)** | **Number (%)** | **Number (%)** |
| Secondary amenorrhea<sup>b</sup> | 2 (9.5%) | 0 (0.0%) | 2 (13.3%) |
| Low T3<sup>c</sup> | 1 (4.8%) | 0 (0.0%) | 1 (6.7%) |
| 1+ high-risk BSI OR 2+ low-risk BSI<sup>d</sup> | 4 (19.0%) | 0 (0.0%) | 4 (26.7%) |
| BMD Z-score < -1<sup>e</sup> | 7 (33.3%) | 0 (0.0%) | 7 (46.7%) |
| Deviation of growth trajectory<sup>f</sup> | NA | NA | NA |
| Elevated EDE-Q score OR clinically diagnosed ED<sup>g</sup> | 3 (14.3%) | 0 (0.0%) | 3 (20.0%) |
| **Secondary Indicators** | **Number (%)** | **Number (%)** | **Number (%)** |
| Oligomenorrhea<sup>h</sup> | 2 (9.5%) | 1 (16.7%) | 1 (6.7%) |
| 1 low-risk BSI | 2 (9.5%) | 0 (0.0%) | 2 (13.3%) |
| Elevated cholesterol<sup>i</sup> | 0 (0.0%) | 0 (0.0%) | 0 (0%) |
| Clinically diagnosed depression and/or anxiety | 4 (19.0%) | 1 (16.7%) | 3 (20.0%) |
| **Emerging Indicators** | **Number (%)** | **Number (%)** | **Number (%)** |
| Iron deficiency | 11 (52.4%) | 2 (33.3%) | 9 (60.0%) |
| Thyroid dysfunction | 4 (19.0%) | 1 (16.7%) | 3 (20.0%) |
| Gastrointestinal health issues | 5 (23.8%) | 0 (0.0%) | 5 (33.3%) |
| Urinary incontinence | 3 (14.3%) | 1 (16.7%) | 2 (13.3%) |

<sup>a</sup> Primary amenorrhea defined as failure to menstruate by age 15 in the presence of normal secondary sexual development.
<sup>b</sup> Secondary amenorrhea defined as absence of 3-11 consecutive menstrual cycles.
<sup>c</sup> Sub-clinically to clinically low total or free T3 defined as within or below the lowest quartile of the reference range (2.0 - 4.4 pg/mL); three participants were excluded from this metric due to use of thyroid medication.
<sup>d</sup> High-risk sites include the femoral neck, sacrum, and pelvis; within the previous 2 years; low-risk sites include all other sites.
<sup>e</sup> For pre-menopausal females, a BMD Z-score less than -1 at the lumbar spine, total hip, or femoral neck.
<sup>f</sup> Not collected.
<sup>g</sup> Elevated global EDE-Q score is defined as >2.30 in females.
<sup>h</sup> Oligomenorrhea defined as >35 days between periods for a maximum of 8 periods/year.
<sup>i</sup> Elevated total or LDL cholesterol is defined as above the reference range; labs missing from one participant in the at-risk group.

##### Statistical Analysis

Data were analysed using IBM SPSS Statistics (v29.0.2.0). Categorical variables were presented as total number and percentages, and continuous variables as mean ± standard deviation for normally distributed ones or median with inter-quartile range (IQR) for non-normally distributed ones. Data was checked for normality using the Shapiro–Wilk test and for equal variance using Levene’s test. Two-sample t-tests were used to compare normally distributed continuous data between the no-risk and at-risk groups. For data that did not meet the assumptions of normality, we used Mann-Whitney U tests to analyse between-group differences. Chi-squared or Fisher’s exact tests were used to compare frequencies of categorical outcomes between groups. Due to the exploratory nature of this study, we relied on internal consistency across multiple measures to draw our conclusion. Therefore, multi-testing correction was not applied to the current statistical significance level of two-sided tests with p<0.05.

## RESULTS

### Participant Characteristics

A total of 21 participants were included in the final analysis (26 ± 3 years old, 164.38 ± 5.44 cm, 59.67 ± 6.48 kg, 22.44 ± 3.02 BMI). Reasons for withdrawal (five participants) included: new medical condition (1), lost to follow-up (2), and birth control usage after the start of the study (2). Demographic information, including age, height, weight, and BMI, grouped by REDs risk group, is presented in Table 2.

**Table 2.**
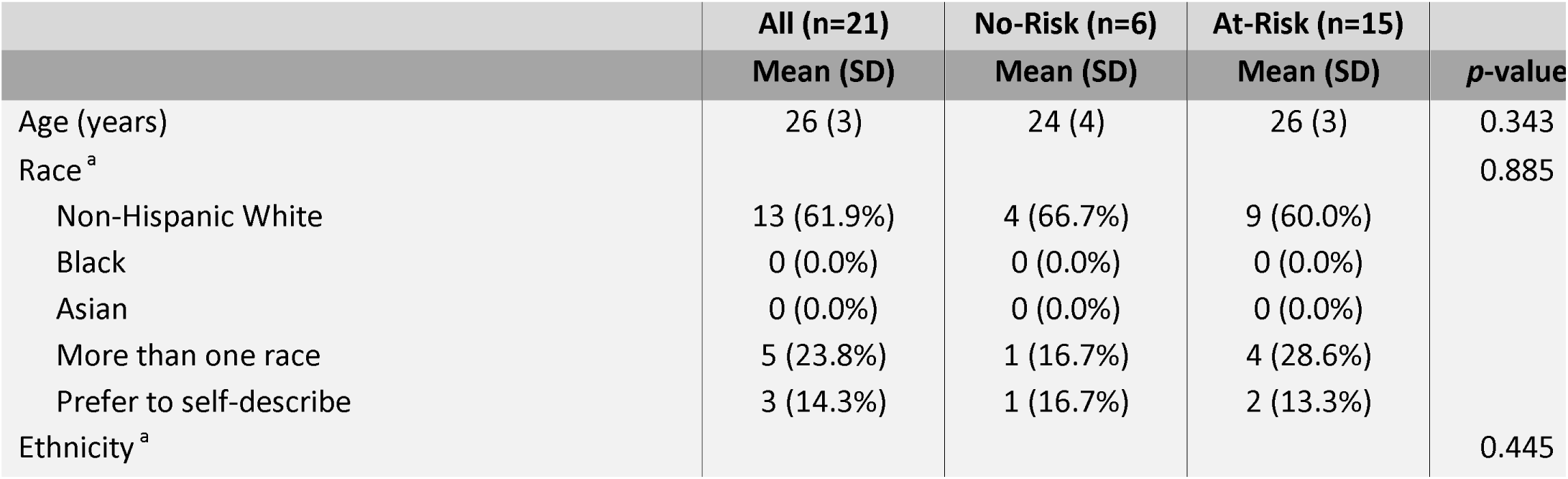
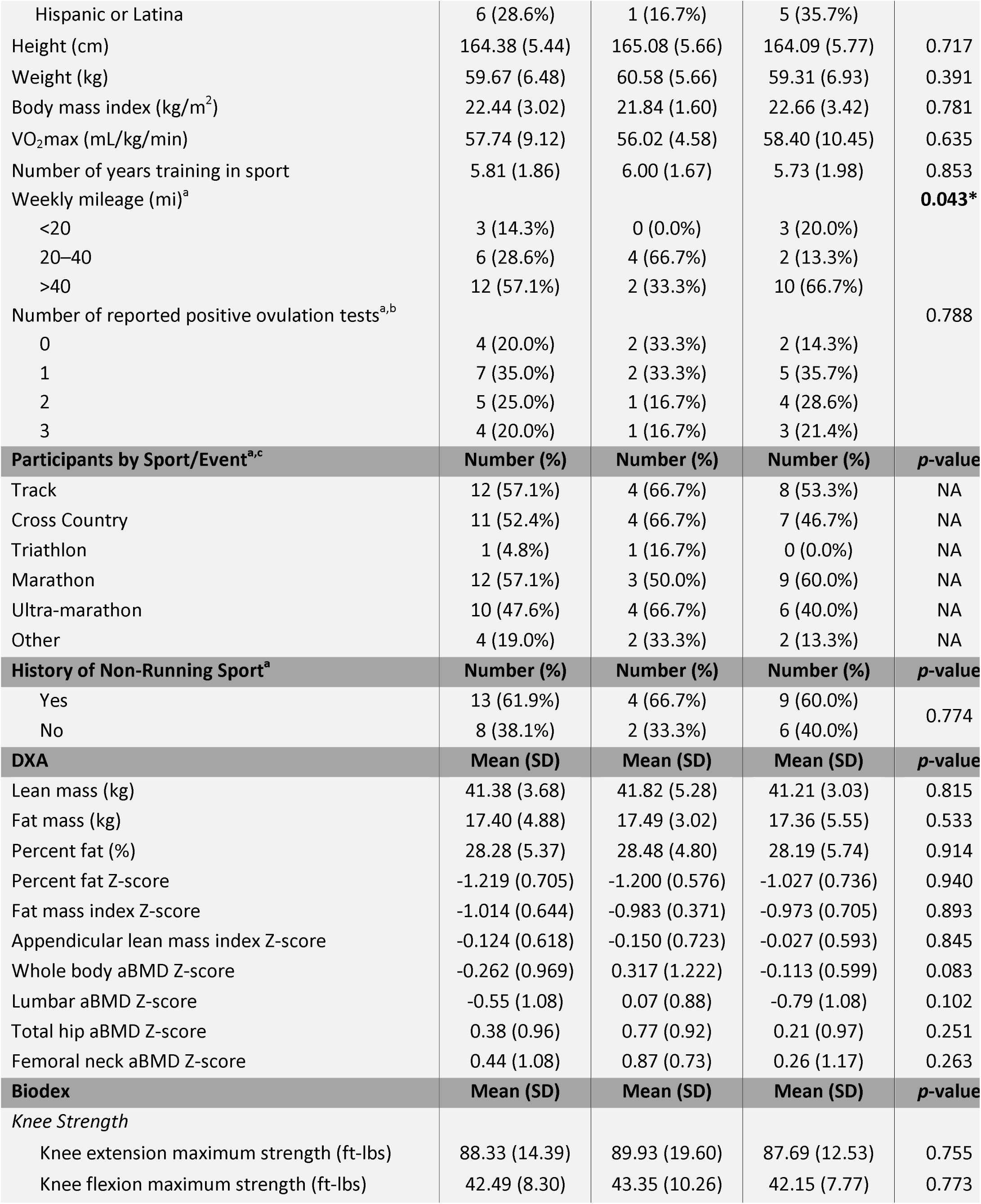
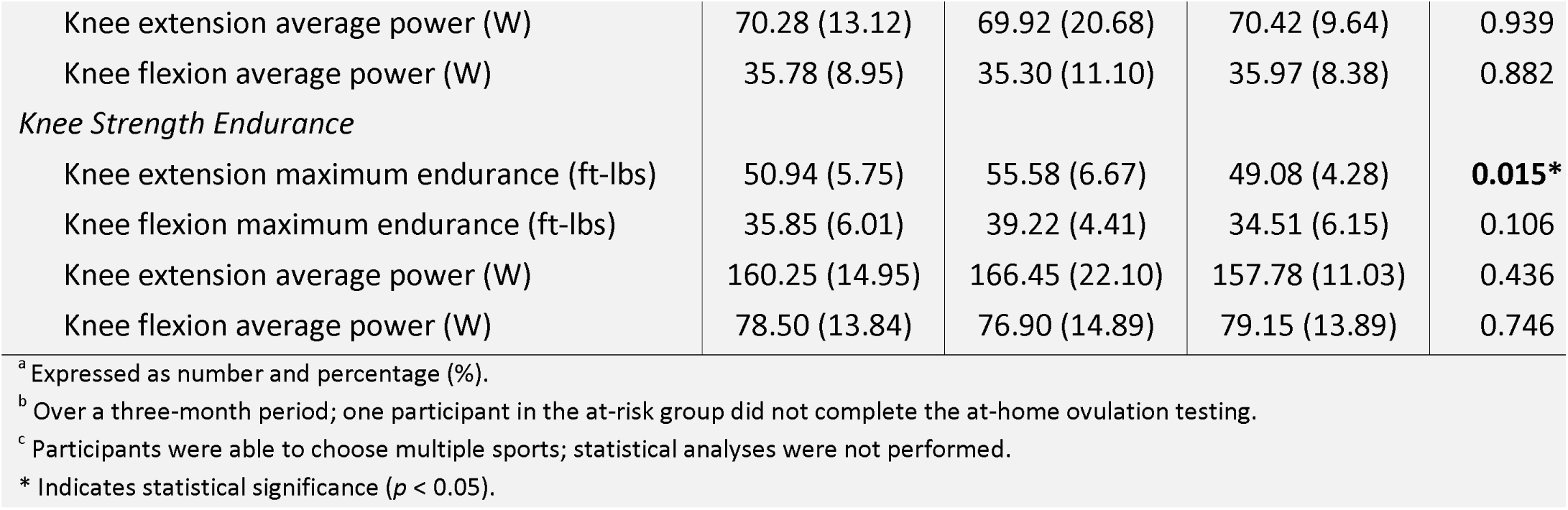
Demographics by REDs Risk.

### REDs Risk Categorization and Clinical Laboratories

Participants were stratified into no-risk (n=6, 28.5%) and at-risk (n=15, 71.5%) groups using the REDs CAT2. Based on the four-color stoplight model, six participants (28.5%) were classified as green (no risk), while the remaining fifteen were classified as at risk: thirteen (62.0%) as yellow, two (9.5%) as orange, and none as red. The prevalence of each REDs CAT2 indicator for this cohort is displayed in Table 1, with low BMD for age (classified as z-score ≤ −1) [25] being the most prevalent indicator in the at-risk group (n=7, 54%). There was a significant difference in reported miles run per week, with a larger proportion of participants in the at-risk group reporting over 40 miles per week (n=10, 66.7% vs. n=2, 33.3%).

Clinical laboratory panels were completed for all but one participant, and only one participant in the at-risk group exhibited sub-clinically low free T3 (Tables 2 and 4). No participants in either group exhibited elevated total or LDL cholesterol (Table 4). Additionally, no participants in the no-risk group reported severe primary or primary indicators of REDs (Table 1).

All but three participants completed the emerging indicators screening via physician interview. The at-risk group reported greater prevalence of iron deficiency (n=9, 60.0% vs. n=2, 33.3%), thyroid dysfunction (n=3, 20.0% vs. n=1, 16.7%), gastrointestinal health issues (n=5, 33.3% vs. n=0, 0.0%), and urinary incontinence (n=2, 13.3% vs. n=1, 16.7%). Additionally, there was no difference between groups in the total number of reported positive ovulation tests (Table 2). Similarly, scores for the EDS-Q and EDE-Q did not differ significantly between groups (Table 4).

### Performance and Nutrition Metrics

VO_2_max did not differ between groups (Table 2), and there was no difference in respiratory quotient, respiratory exchange ratio, or anaerobic threshold (Table 4). Notably, maximum extensor strength was significantly less in the at-risk group than the no-risk group (49.08 ± 4.28, 55.58 ± 6.67, respectively, p=0.015). All other metrics from the Biodex assessments were not significantly different (Table 2).

There was no difference in total caloric or macronutrient, cholesterol, dietary fiber, iron, or calcium intake between groups (Table 4).

### DXA and HR-pQCT Measures

There was no significant difference between groups in mean values for all DXA and HR-pQCT variables (Table 2 and 3 for z-scores, raw values in Table 4); however, 12 out of 14 HR-pQCT metrics trended worse in the at-risk group (Figure 1). One participant was unable to complete the total hip and femoral neck DXA scans due to a history of bilateral BSIs. Seven participants had low BMD for age (defined as z-score < −1.0) at the lumbar spine, femoral neck, or total hip, and all were in the at-risk group. In addition, three of these participants had BMD Z-scores of ≤ −2.0.

**Figure 1.**
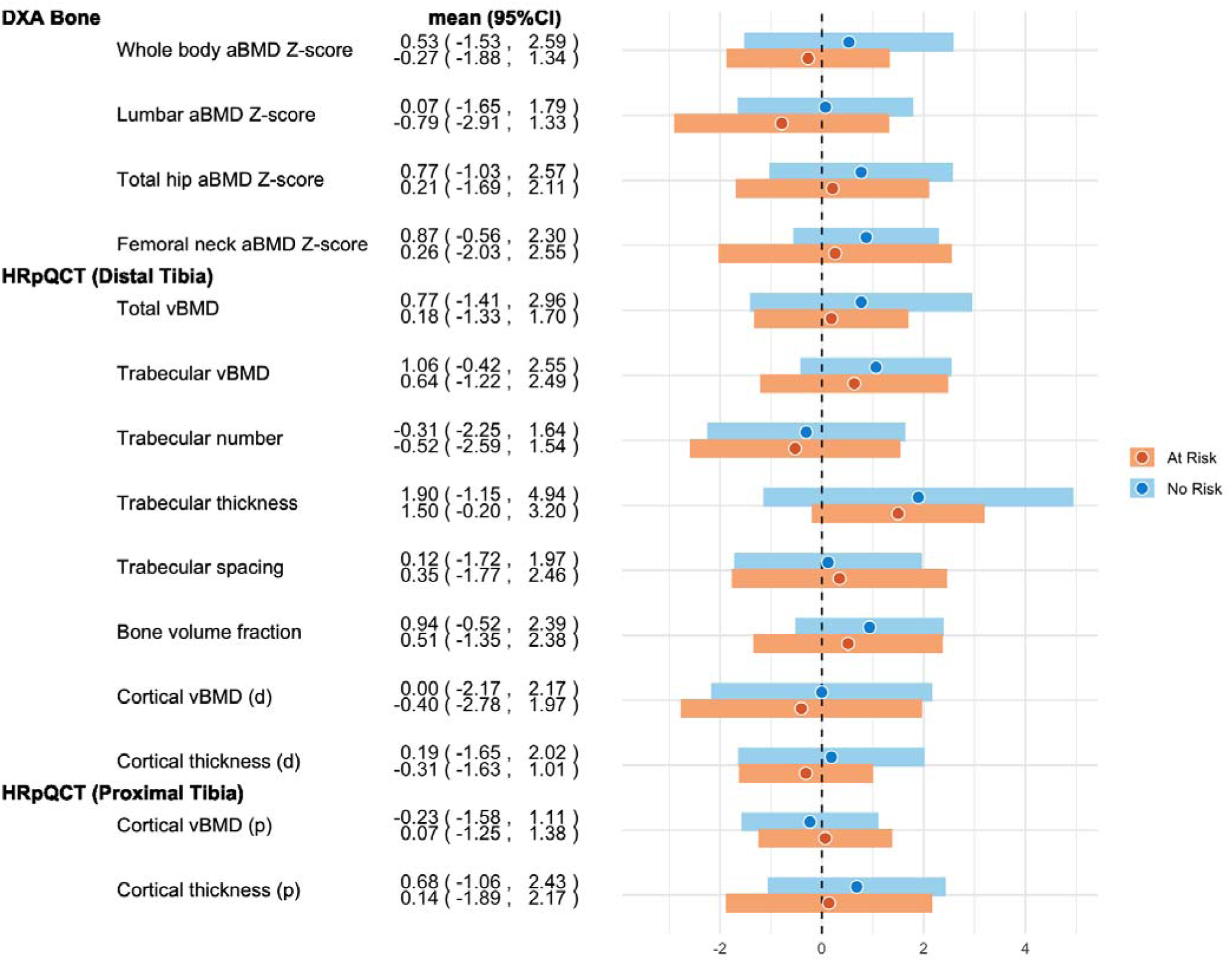
Forest Plot for DXA and HRpQCT Bone Measurement Z-scores Between At-Risk and No-Risk Groups

**Table 3.** HR-pQCT Results for No-Risk and At-Risk Groups.

|  | No-Risk (n=6) | At-Risk (n=15) |  |
| --- | --- | --- | --- |
| Metaphyseal Tibia | Mean (SD) | Mean (SD) | p-value |
| *Volumetric BMD Z-scores* |  |  |  |
| Total BMD | 0.772 (1.115) | 0.185 (0.775) | 0.182 |
| Cortical BMD | -0.003 (1.108) | -0.403 (1.211) | 0.312 |
| Trabecular BMD | 1.063 (0.757) | 0.635 (0.944) | 0.213 |
| *Geometry Z-scores* |  |  |  |
| Total area | -0.319 (0.972) | -0.374 (0.784) | 0.894 |
| Cortical area | -0.004 (0.623) | -0.502 (0.719) | 0.154 |
| *Microarchitecture Z-scores* |  |  |  |
| Cortical thickness | 0.186 (0.935) | -0.313 (0.673) | 0.185 |
| Trabecular number | -0.307 (0.993) | -0.525 (1.056) | 0.668 |
| Trabecular thickness | 1.896 (1.555) | 1.499 (0.867) | 0.460 |
| Trabecular spacing | 0.122 (0.942) | 0.346 (1.080) | 0.663 |
| Bone volume fraction | 0.936 (0.743) | 0.514 (0.950) | 0.161 |
| **Diaphyseal Tibia** | **Mean (SD)** | **Mean (SD)** | **p-value** |
| *Volumetric BMD Z-score* |  |  |  |
| Cortical BMD | -0.234 (0.687) | 0.066 (0.672) | 0.312 |
| *Geometry Z-scores* |  |  |  |
| Total area | 0.105 (0.531) | -0.073 (0.653) | 0.562 |
| Cortical area | 0.333 (0.440) | -0.032 (0.822) | 0.320 |
| *Microarchitecture Z-score* |  |  |  |
| Cortical thickness | 0.684 (0.892) | 0.139 (1.035) | 0.273 |

## DISCUSSION

Although we did not detect statistically significant differences between groups within our HR-pQCT parameters, our findings consistently trended in the hypothesized direction: athletes at risk for REDs demonstrated poorer bone geometry, microarchitecture, and BMD than their no-risk counterparts. The lack of statistically significant differences in absolute bone mineral density values between groups was expected, given the small sample size. The use of z-scores for HR-pQCT provided additional clinical insight and helped to identify meaningful patterns not captured by absolute values alone. As a pilot study, these findings serve as a foundation for future research in larger, adequately powered cohorts.

Several variables that were notably worse in the at-risk group included cortical area and cortical thickness at the metaphyseal tibia, as well as cortical thickness and cortical porosity at the diaphyseal tibia. These trends are consistent with findings from prior studies evaluating bone quality in individuals with indicators of LEA across a spectrum of severity. For example, in a study of winter endurance athletes, cortical area at the radius was significantly lower in the at-risk group compared to low-risk counterparts [19]. These trends demonstrate the potential of HR-pQCT as a clinically meaningful tool for assessing REDs risk.

### Clinical Implications

Approximately one-third of participants demonstrated low bone mineral density (at lumbar spine, femoral neck, or total hip), and all those individuals were classified in the at-risk group. In addition, 28% of participants reported a history of bone stress injury, all of whom were also in the at-risk group. Prior research has similarly shown increased prevalence of high-risk or recurrent BSIs among athletes exhibiting markers of LEA [26,27]. A recent prospective study further highlighted this relationship, reporting a 25-fold greater likelihood of high-risk BSI within the previous two years and nearly an 8-fold greater risk of new BSI in athletes classified in the REDs orange risk category compared to those in the green risk group [28]. These findings support the inclusion of bone health history as a critical marker of REDs risk and emphasize the need for early screening and monitoring among endurance athletes.

The high prevalence of primary or secondary amenorrhea (n=5, 23.8%), low BMD (at lumbar spine, femoral neck, and total hip, n=7, 33.3%), and BSI history (n=4, 19.0%) observed in the at-risk group aligns with decades of research linking these outcomes to LEA [25,29–32]. Of interest, only one participant (in the at-risk group) exhibited clinically low levels of free T3. Suppressed thyroid function, specifically reductions in T3, is a well-documented physiological consequence of low energy availability. However, in a previous study [28], low or low-normal free T3 was only observed in individuals at moderate to high REDs risk (orange and red categories). Given that only two participants in our cohort were classified as orange risk, it is likely that the severity or chronicity of LEA in this sample was insufficient to result in significant thyroid suppression.

One-third of all participants (n=7, 33.3%) reported a history of clinically diagnosed depression, anxiety, or eating disorders, or demonstrated elevated scores on the EDE-Q. These findings are notable given the established association between psychological conditions, particularly depressive symptoms, and REDs [33]. Routine screening for mental health concerns is essential when evaluating athletes for REDs risk.

Reflecting patterns observed in previous literature, participants in the at-risk group reported higher rates of iron deficiency history [34,35], thyroid dysfunction [36–40], and gastrointestinal (GI) issues [41,42] compared to those in the no-risk group. While these emerging indicators are less well-established in the literature, our findings highlight the importance of research in these spaces to better understand their potential role in identifying or contributing to REDs risk.

### Limitations

As a pilot study, it was inherently underpowered with small and uneven group sizes. The cross-sectional design for the imaging limited causal interpretation. Additionally, strict inclusion criteria, such as requiring participants to be currently uninjured and not using hormonal contraceptives, may have excluded individuals at higher REDs risk. Finally, self-reported nutritional intake data are subject to recall bias and may not accurately reflect true energy availability.

Despite these limitations, this study represents a meaningful step toward understanding the effects of REDs on bone geometry and microarchitecture in female endurance athletes. The integration of HR-pQCT imaging with REDs-CAT2 screening may allow for a richer characterization of REDs-related bone health than DXA alone. These preliminary findings underscore the importance of continued research on REDs risk in female endurance athletes.

## CONCLUSION

In conclusion, while statistical significance was not achieved, consistent trends in HR-pQCT outcomes suggest a physiological pattern linking REDs risk with compromised bone health. In future studies, integrating z-score-based bone health profiling with longitudinal assessments in a larger cohort of diverse risk profiles may provide a more nuanced picture of REDs-related bone health decline or resilience.

## Data Availability

All data relevant to the study are included in the article or uploaded as online supplemental material. Additional data are available upon reasonable request.

## Funding Section or Acknowledgements

Thank you to Stanford’s Women’s Health & Sex Differences in Medicine Center for funding this study, and to Wu Tsai Human Performance Alliance for supporting the postdoctoral fellowship for the project.

## Contributors

All authors made substantial contributions to the manuscript. MS, JF, and EK conceptualized, drafted, and revised the manuscript, with MS and JF making equal contributions to the work. KK and JL contributed to writing and revising the article and drafting the figures. JL and YZ supported data management and analysis. AR and MPS provided feedback on the final manuscript draft. Yair Blumberg conducted the VO_2_max testing on participants.

## Patient and Public Involvement

Patients and the public were not involved in the design, conduct, reporting, or dissemination of this research.

## Ethical Approval

This study involves human participants and was approved by the Institutional Review Board of Stanford University School of Medicine (IRB #69308). Participants gave informed consent prior to participating in the study.

## Equity, Diversity, and Inclusion

Our study was on women runners in the San Francisco Bay Area, USA. We did not purposefully recruit people from marginalized communities. Our author team included two men and six women (first and senior author are women). The authors’ disciplines include exercise science, sports medicine, musculoskeletal imaging, genetics, statistics, and human biology, and we included two junior scholars. While our study focused on female athletes, we did not examine the effects of race/ethnicity or socioeconomic status.

## References

1. Hutson MJ, O’Donnell E, Brooke-Wavell K, Sale C, Blagrove RC. Effects of Low Energy Availability on Bone Health in Endurance Athletes and High-Impact Exercise as A Potential Countermeasure: A Narrative Review. Sports Med Auckl Nz. 2021;51(3):391–403.

2. Indirli R, Lanzi V, Mantovani G, Arosio M, Ferrante E. Bone health in functional hypothalamic amenorrhea: What the endocrinologist needs to know. Front Endocrinol. 2022 Oct 11;13:946695.

3. Schuit SCE, van der Klift M, Weel AEAM, de Laet CEDH, Burger H, Seeman E, et al. Fracture incidence and association with bone mineral density in elderly men and women: the Rotterdam Study. Bone. 2004 Jan 1;34(1):195–202.

4. Trajanoska K, Schoufour JD, de Jonge EAL, Kieboom BCT, Mulder M, Stricker BH, et al. Fracture incidence and secular trends between 1989 and 2013 in a population based cohort: The Rotterdam Study. Bone. 2018 Sept 1;114:116–24.

5. Melton LJ III, Crowson CS, O’Fallon WM, Wahner HW, Riggs BL. Relative Contributions of Bone Density, Bone Turnover, and Clinical Risk Factors to Long-Term Fracture Prediction1*. J Bone Miner Res. 2003 Feb 1;18(2):312–8.

6. Bouxsein ML, Karasik D. Bone geometry and skeletal fragility. Curr Osteoporos Rep. 2006 June 1;4(2):49–56.

7. Sarfati M, Chapurlat R, Dufour AB, Sornay-Rendu E, Merle B, Boyd SK, et al. Short-term risk of fracture is increased by deficits in cortical and trabecular bone microarchitecture independent of DXA BMD and FRAX: Bone Microarchitecture International Consortium (BoMIC) prospective cohorts. J Bone Miner Res Off J Am Soc Bone Miner Res. 2024 Oct 29;39(11):1574–83.

8. Elizabeth J. S, Broe KE, Xu H, Yang L, Boyd S, Biver E, et al. Cortical and trabecular bone microarchitecture predicts incident fracture independently of DXA bone mineral density and FRAX in older women and men: The Bone Microarchitecture International Consortium (BoMIC). Lancet Diabetes Endocrinol. 2019 Jan;7(1):34–43.

9. Sornay-Rendu E, Duboeuf F, Chapurlat RD. Postmenopausal women with normal BMD who have fractures have deteriorated bone microarchitecture: A prospective analysis from The OFELY study. Bone. 2024 May 1;182:117072.

10. Tenforde AS, Kraus E, Fredericson M. Bone Stress Injuries in Runners. Phys Med Rehabil Clin N Am. 2016 Feb 1;27(1):139–49.

11. Tenforde AS, Ackerman KE, Bouxsein ML, Gaudette L, McCall L, Rudolph SE, et al. Factors Associated With High-Risk and Low-Risk Bone Stress Injury in Female Runners: Implications for Risk Factor Stratification and Management. Orthop J Sports Med. 2024 May 21;12(5):23259671241246227.

12. Mountjoy M, Sundgot-Borgen J, Burke L, Carter S, Constantini N, Lebrun C, et al. The IOC consensus statement: beyond the Female Athlete Triad—Relative Energy Deficiency in Sport (RED-S). 2014 Apr 1 [cited 2025 Apr 15]; Available from: https://bjsm.bmj.com/content/48/7/491.long

13. Papageorgiou M, Dolan E, Elliott-Sale KJ, Sale C. Reduced energy availability: implications for bone health in physically active populations. Eur J Nutr. 2018;57(3):847–59.

14. Medicine BPGL and BA of S and E. International olympic committee relative energy deficiency in sport clinical assessment tool 2 (IOC REDs CAT2). 2023 Sept 1 [cited 2025 Apr 15]; Available from: https://bjsm.bmj.com/content/57/17/1068

15. Schipilow JD, Macdonald HM, Liphardt AM, Kan M, Boyd SK. Bone micro-architecture, estimated bone strength, and the muscle-bone interaction in elite athletes: An HR-pQCT study. Bone. 2013 Oct 1;56(2):281–9.

16. Bolotin HH, Sievänen H, Grashuis JL, Kuiper JW, Järvinen TLN. Inaccuracies Inherent in Patient-Specific Dual-Energy X-Ray Absorptiometry Bone Mineral Density Measurements: Comprehensive Phantom-Based Evaluation. J Bone Miner Res. 2001;16(2):417–26.

17. Garg MK, Kharb S. Dual energy X-ray absorptiometry: Pitfalls in measurement and interpretation of bone mineral density. Indian J Endocrinol Metab. 2013;17(2):203–10.

18. Bolotin HH. Inaccuracies inherent in dual-energy X-ray absorptiometry in vivo bone mineral densitometry may flaw osteopenic/osteoporotic interpretations and mislead assessment of antiresorptive therapy effectiveness. Bone. 2001 May 1;28(5):548–55.

19. Wyatt PM, Drager K, Groves EM, Stellingwerff T, Billington EO, Boyd SK, et al. Comparison of Bone Quality Among Winter Endurance Athletes with and Without Risk Factors for Relative Energy Deficiency in Sport (REDs): A Cross-Sectional Study. Calcif Tissue Int. 2023 Oct 1;113(4):403–15.

20. Ackerman KE, Nazem T, Chapko D, Russell M, Mendes N, Taylor AP, et al. Bone Microarchitecture Is Impaired in Adolescent Amenorrheic Athletes Compared with Eumenorrheic Athletes and Nonathletic Controls. J Clin Endocrinol Metab. 2011 Oct;96(10):3123–33.

21. Harris PA, Taylor R, Minor BL, Elliott V, Fernandez M, O’Neal L, et al. The REDCap Consortium: Building an International Community of Software Platform Partners. J Biomed Inform. 2019 July;95:103208.

22. Harris PA, Taylor R, Thielke R, Payne J, Gonzalez N, Conde JG. Research Electronic Data Capture (REDCap) - A metadata-driven methodology and workflow process for providing translational research informatics support. J Biomed Inform. 2009 Apr;42(2):377–81.

23. O’Donnell J, McCluskey P, Stellingwerff T. Ovulation Monitoring Protocol [Internet]. 2022 [cited 2025 Aug 5]. Available from: http://www.csipacific.ca/wp-content/uploads/2022/09/CSI-Pacific-Ovuation-Protocol-20220921.pdf

24. Smith EM, Drager K, Groves EM, Gabel L, Boyd SK, Burt LA. New approach to identifying elite winter sport athletes’ risk of relative energy deficiency in sport (REDs). BMJ Open Sport Exerc Med [Internet]. 2025 Jan 30 [cited 2025 Apr 24];11(1). Available from: https://bmjopensem.bmj.com/content/11/1/e002320

25. Souza MJD, Nattiv A, Joy E, Misra M, Williams NI, Mallinson RJ, et al. 2014 Female Athlete Triad Coalition Consensus Statement on Treatment and Return to Play of the Female Athlete Triad: 1st International Conference held in San Francisco, California, May 2012 and 2nd International Conference held in Indianapolis, Indiana, May 2013. 2014 Feb 1 [cited 2025 July 21]; Available from: https://bjsm.bmj.com/content/48/4/289.long

26. Roche M, Nattiv A, Sainani K, Barrack M, Kraus E, Tenforde A, et al. Higher Triad Risk Scores Are Associated With Increased Risk for Trabecular-Rich Bone Stress Injuries in Female Runners. Clin J Sport Med. 2023 Nov;33(6):631.

27. Gehman S, Ackerman KE, Caksa S, Rudolph SE, Hughes JM, Garrahan M, et al. Restrictive Eating and Prior Low-Energy Fractures Are Associated With History of Multiple Bone Stress Injuries. 2022 May 6 [cited 2025 July 21]; Available from: https://journals.humankinetics.com/view/journals/ijsnem/32/5/article-p325.xml

28. Heikura IA, McCluskey WTP, Tsai MC, Johnson L, Murray H, Mountjoy M, et al. Application of the IOC Relative Energy Deficiency in Sport (REDs) Clinical Assessment Tool version 2 (CAT2) across 200+ elite athletes. Br J Sports Med [Internet]. 2024 Aug 20 [cited 2024 Oct 15]; Available from: https://bjsm.bmj.com/content/early/2024/08/19/bjsports-2024-108121

29. Cobb KL, Bachrach LK, Greendale G, Marcus R, Neer RM, Nieves J, et al. Disordered Eating, Menstrual Irregularity, and Bone Mineral Density in Female Runners. Med Sci Sports Exerc. 2003 May;35(5):711.

30. Gibbs JC, Nattiv A, Barrack MT, Williams NI, Rauh MJ, Nichols JF, et al. Low Bone Density Risk Is Higher in Exercising Women with Multiple Triad Risk Factors. Med Sci Sports Exerc. 2014 Jan;46(1):167.

31. Nose-Ogura S, Yoshino O, Dohi M, Kigawa M, Harada M, Kawahara T, et al. Low Bone Mineral Density in Elite Female Athletes With a History of Secondary Amenorrhea in Their Teens. Clin J Sport Med Off J Can Acad Sport Med. 2020 May;30(3):245–50.

32. Ackerman KE, Cano Sokoloff N, Maffazioli GDN, Clarke H, Lee H, Misra M. Fractures in Relation to Menstrual Status and Bone Parameters in Young Athletes. Med Sci Sports Exerc. 2015 Aug;47(8):1577–86.

33. Pensgaard AM, Sundgot-Borgen J, Edwards C, Jacobsen AU, Mountjoy M. Intersection of mental health issues and Relative Energy Deficiency in Sport (REDs): a narrative review by a subgroup of the IOC consensus on REDs. 2023 Sept 1 [cited 2025 July 21]; Available from: https://bjsm.bmj.com/content/57/17/1127

34. McKay AKA, Pyne DB, Burke LM, Peeling P. Iron Metabolism: Interactions with Energy and Carbohydrate Availability. Nutrients. 2020 Nov 30;12(12):3692.

35. Finn EE, Tenforde AS, Fredericson M, Golden NH, Carson TL, Karvonen-Gutierrez CA, et al. Markers of Low-Iron Status Are Associated with Female Athlete Triad Risk Factors. Med Sci Sports Exerc. 2021 Sept;53(9):1969.

36. Areta JL, Taylor HL, Koehler K. Low energy availability: history, definition and evidence of its endocrine, metabolic and physiological effects in prospective studies in females and males. Eur J Appl Physiol. 2021;121(1):1–21.

37. Skrzypiec-Spring M, Kuliczkowska-Płaksej J, Szeląg A, Bolanowski M. Atypical thyroid tests in an athlete treated for hypothyroidism as the first symptom of pituitary dysfunction due to relative energy deficiency. Endocrinol Diabetes Metab Case Rep. 2024 Oct 29;2024(4):24–0066.

38. McCall LM, Ackerman KE. Endocrine and metabolic repercussions of relative energy deficiency in sport. Curr Opin Endocr Metab Res. 2019 Dec 1;9:56–65.

39. Loucks AB, Verdun M, Heath EM. Low energy availability, not stress of exercise, alters LH pulsatility in exercising women. J Appl Physiol. 1998 Jan;84(1):37–46.

40. Mathisen TF, Heia J, Raustøl M, Sandeggen M, Fjellestad I, Sundgot-Borgen J. Physical health and symptoms of relative energy deficiency in female fitness athletes. Scand J Med Sci Sports. 2020;30(1):135–47.

41. Ackerman KE, Holtzman B, Cooper KM, Flynn EF, Bruinvels G, Tenforde AS, et al. Low energy availability surrogates correlate with health and performance consequences of Relative Energy Deficiency in Sport. 2019 May 1 [cited 2025 July 22]; Available from: https://bjsm.bmj.com/content/53/10/628?ijkey=9dad608fe943dc88b07df96e77d62ddeb218e39f&keytype2=tf_ipsecsha

42. Kuikman MA, Mountjoy M, Burr JF. Examining the Relationship between Exercise Dependence, Disordered Eating, and Low Energy Availability. Nutrients. 2021 Aug;13(8):2601.

