## Supplementary material for "Comparison of DXA and HR-pQCT Measures Among Female Runners at Risk for Relative Energy Deficiency in Sport (REDs): A Pilot Study": Table 4. Supplemental Data

| **Table 4.** Supplemental Data | | | | | |
| --- | --- | --- | --- | --- | --- |
|  | **No-Risk (n=6)** | | **At-Risk (n=15)** | |  |
| **Validated Surveys** | **Median** | **IQR** | **Median** | **IQR** | ***p*-value** |
| EDE-Q Global Score | 1.29 | 1.19 – 1.34 | 1.54 | 1.19 – 1.88 | 0.257 |
| EDS Total Score | 53 | 46 – 58 | 53 | 42 – 72 | 0.476 |
| **Nutritional Intake** | **Median** | **IQR** | **Median** | **IQR** | ***p*-value** |
| Energy (kcal) | 2120.94 | 1868.31 – 2752.93 | 2030.55 | 1541.28 – 2634.65 | 0.655 |
| Protein (g) | 96.03 | 88.77 – 120.82 | 86.44 | 75.44 – 133.94 | 0.618 |
| Fat (g) | 88.07 | 74.24 – 119.66 | 81.98 | 61.08 – 113.98 | 0.530 |
| Carbohydrate (g) | 247.37 | 202.24 – 300.37 | 242.00 | 171.87 – 313.16 | 0.877 |
| Dietary fiber (g) | 27.57 | 25.10 – 31.81 | 27.57 | 19.62 – 41.71 | 0.934 |
| Iron (mg) | 17.88 | 16.09 – 19.74 | 18.20 | 11.50 – 22.78 | 0.869 |
| Calcium (mg) | 1155.35 | 1037.36 – 1518.35 | 1275.88 | 692.45 – 2055.68 | 0.998 |
| **Lab Values** | **Median** | **IQR** | **Median** | **IQR** | ***p*-value** |
| Total cholesterol | 153 | 144 – 164 | 159 | 151 – 173 | 0.579 |
| HDL cholesterol | 59 | 53 – 71 | 70 | 64 – 83 | 0.108 |
| Triglycerides | 73 | 65 – 82 | 60 | 43 – 68 | **0.035*** |
| Free T3 | 3.5 | 3.2 – 3.6 | 3.2 | 2.8 – 3.2 | 0.106 |
| **Performance Data** | **Median** | **IQR** | **Median** | **IQR** | ***p*-value** |
| VO2 max (mL/min/kg) | 55.1 | 53.5 – 60.6 | 58.0 | 53.5 – 63.4 | 0.635 |
| Respiratory Quotient | 1.02 | 1.00 – 1.04 | 1.09 | 1.05 – 1.10 | 0.137 |
| Peak RER | 1.01 | 0.98 – 1.04 | 1.09 | 1.05 – 1.11 | 0.053 |
| Anaerobic Threshold | 29.6 | 28.0 – 35.0 | 30.5 | 27.1 – 41.1 | 0.327 |
| **DXA Raw Values** | **Median** | **IQR** | **Median** | **IQR** | ***p*-value** |
| Whole body BMD | 1.123 | 1.103 – 1.160 | 1.090 | 1.056 – 1.120 | 0.099 |
| Lumbar spine BMD | 1.016 | 0.998 – 1.063 | 0.962 | 0.882 – 1.007 | 0.148 |
| Total hip BMD | 1.004 | 0.976 – 1.045 | 0.935 | 0.883 – 1.033 | 0.241 |
| Femoral neck BMD | 0.941 | 0.900 – 0.969 | 0.875 | 0.774 – 0.946 | 0.236 |
| Total percent fat (%) | 26.7 | 25.2 – 32.7 | 26.3 | 23.6 – 30.6 | 0.914 |
| Fat mass index | 5.33 | 5.14 – 7.28 | 5.63 | 4.22 – 7.04 | 0.533 |
| Appendicular lean mass index | 6.46 | 6.00 – 7.31 | 6.40 | 6.06 – 6.98 | 0.850 |
| **HR-pQCT Raw Values** | **Median** | **IQR** | **Median** | **IQR** | ***p*-value** |
| *Metaphyseal Tibia - Density* |  |  |  |  |  |
| Total vBMD (mg HA/cm^3^) | 353.50 | 318.15 - 418.45 | 338.70 | 299.40 - 361.40 | 0.161 |
| Trabecular vBMD (mg HA/cm^3^) | 213.25 | 196.53 - 254.20 | 199.00 | 181.50 - 221.60 | 0.186 |
| Cortical vBMD (mg HA/cm^3^) | 944.60 | 908.70 - 985.35 | 913.70 | 896.80 - 968.60 | 0.436 |
| Cortical TMD (mg HA/cm^3^) | 1047.00 | 995.90 - 1071.78 | 1036.60 | 1033.20 - 1063.80 | 0.748 |
| *Metaphyseal Tibia - Geometry* |  |  |  |  |  |
| Total area (mm^2^) | 670.50 | 569.70 - 702.05 | 636.70 | 610.20 - 688.70 | 0.837 |
| Trabecular area (mm^2^) | 546.15 | 444.35 - 593.3 | 522.70 | 507.30 - 585.00 | 0.924 |
| Cortical area (mm^2^) | 121.90 | 114.15 - 141.575 | 109.00 | 103.40 - 132.00 | 0.152 |
| *Metaphyseal Tibia - Microarchitecture* |  |  |  |  |  |
| Bone volume fraction | 0.308 | 0.283 - 0.353 | 0.280 | 0.257 - 0.316 | 0.161 |
| Trabecular number (1/mm) | 1.408 | 1.242 - 1.408 | 1.312 | 1.202 - 1.484 | 0.635 |
| Trabecular thickness (mm) | 0.291 | 0.274 - 0.324 | 0.284 | 0.281 - 0.298 | 0.389 |
| Trabecular spacing (mm) | 0.660 | 0.555 - 0.736 | 0.697 | 0.621 - 0.776 | 0.577 |
| Cortical thickness (mm) | 1.465 | 1.300 - 1.698 | 1.274 | 1.240 - 1.557 | 0.182 |
| Cortical porosity (%) | 0.009 | 0.008 - 0.015 | 0.019 | 0.008 - 0.031 | 0.310 |
| *Diaphyseal Tibia - Density* |  |  |  |  |  |
| Total vBMD (mg HA/cm^3^) | 812.40 | 780.03 - 868.20 | 812.00 | 791.50 - 853.20 | 0.727 |
| Cortical vBMD (mg/cm^3^) | 1048.15 | 1028.07 - 1053.80 | 1054.80 | 1036.00 - 1061.10 | 0.392 |
| Cortical TMD (mg HA/cm^3^) | 1073.85 | 1052.07 - 1077.92 | 1077.60 | 1060.90 - 1090.40 | 0.350 |
| *Diaphyseal Tibia - Geometry* |  |  |  |  |  |
| Total area (mm^2^) | 352.00 | 316.40 - 366.95 | 334.40 | 329.40 - 358.40 | 0.556 |
| Cortical area (mm^2^) | 265.85 | 255.825 - 280.975 | 255.50 | 228.90 - 285.30 | 0.323 |
| *Diaphyseal Tibia - Microarchitecture* |  |  |  |  |  |
| Cortical porosity (%) | 0.007 | 0.003 - 0.011 | 0.008 | 0.007 - 0.015 | 0.304 |
| Cortical thickness (mm) | 5.916 | 5.625 - 6.948 | 5.814 | 5.312 - 6.478 | 0.297 |
